# The Growth Laws of Brain Metastases

**DOI:** 10.1101/2022.02.03.22270146

**Authors:** Beatriz Ocaña-Tienda, Julián Pérez-Beteta, Juan Jiménez-Sánchez, David Molina García, Ana Ortiz de Mendivil, Beatriz Asenjo, David Albillo, Luís A. Pérez-Romasanta, Manuel Valiente, Lucía Zhu, Pedro García-Gómez, Elisabet González-Del Portillo, Manuel Llorente, Natalia Carballo, Estanislao Arana, Víctor M. Pérez-García

## Abstract

Tumor growth is the result of the interplay of complex biological processes in huge numbers of individual cells living in changing environments. Effective simple mathematical laws have been shown to describe tumor growth in vitro, or simple animal models with bounded-growth dynamics accurately. However, results for the growth of human cancers in patients are scarce. Our study mined a large dataset of 1133 brain metastases (BMs) with longitudinal imaging follow-up to find growth laws for untreated BMs and recurrent treated BMs. Untreated BMs showed high growth exponents, most likely related to the underlying evolutionary dynamics, with experimental tumors in mice resembling accurately the disease. Recurrent BMs growth exponents were smaller, most probably due to a reduction in tumor heterogeneity after treatment, which may limit the tumor evolutionary capabilities. In silico simulations using a stochastic discrete mesoscopic model with basic evolutionary dynamics led to results in line with the observed data.

## Introduction

Macroscopic tumor growth is a complex process resulting from the interplay of different biological elements at the cellular and subcellular levels. These include the driving molecular alterations and their associated heterogeneity, angiogenesis, the immune system, the tumor microenvironment, and surrounding healthy structures, the effect of treatments on the different tumor phenotype-s/genotypes, etc.

Mathematical growth laws have been shown to describe longitudinal tumor growth dynamics effectively in simple experimental models. [1–5]. A great deal of data is available for those models, which do not have the biological complexity of human tumors. One would expect that describing cancer growth mathematically in humans would be far more difficult because of the different biological mechanisms that drive it over distinct tumor stages.

Assessing complex tumor growth dynamics over time, given the large genotypic and phenotypic variability developed during tumorigenesis, is very difficult with current techniques. Medical images are performed routinely in most cancer patients and provide rough global macroscopic information - the so-called imaging phenotype-which integrates the several processes occurring at the microscale, potentially providing information on the underlying tumor biology.

Longitudinal datasets for untreated malignant tumors are rare, and of limited quality, since therapeutic action is typically performed promptly. This is why studies of untreated tumor gynamics laws in humans have been mostly limited to the use of two time points per patient what allows only for the indentification of average growth rates [6, 7]. Only recently has a study using more time points shown explosive dynamics, argued to be the result of evolutionary dynamics in human cancers [8].

Our goal in this study combining clinical data and mathematical models, was to provide more evidences on the macroscopic growth dynamics of both treated and untreated tumors in human patients. We focused on growth dynamics of brain metastases (BMs) integrating mathematical and computational models with data analysis methods on a patient database with longitudinal MR imaging follow-up and more than a thousand lesions. BMs are the most common intracranial tumor and a major complication of many cancers, with 20%-30% of cancer patients developing the condition in the course of their disease [9, 10]. Here we focused on two scenarios. The first one was the growth of untreated BMs. Human beings as well as mice data were studied. The second scenario considered BMs under different treatment modalities. Therapy options for BMs are surgery, whole brain radiation therapy (WBRT), gamma knife radiosurgery, stereotactic radiosurgery (SRS), targeted and systemic therapies.

We wanted to investigate whether there was a measurable effect on the growth dynamics of the expected loss of tumor heterogeneity after treatment. To this end, we performed an analysis of the growth dynamics of untreated BMs, and BMs treated under different types of therapy, by fitting a general tumor growth law [11]. The results obtained were endorsed by *in silico* simulations with a discrete stochastic model of tumor growth, which helped to gain insight into the causes of the behavior observed in BMs.

## Results

### Brain metastasis growth is explosive in untreated patients but not in treated ones

The Von-Bertalanffy model [11]

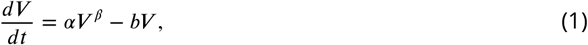

has been proven recently to describe the longitudinal dynamics of growing human tumors [8]. It has been argued, and supported with data from different human cancers, that malignant tumors with heterogeneous clonal composition have exponents *β* > 1. The reason is that in heterogeneous tumors there would be a range of phenotype/genotypes to choose from, leading to selection for more aggressive phenotypes/genotypes, and an acceleration of the growth rate, which would manifest in the form of faster-than-exponential explosive unbounded growth. In this paper will focus on scenarios of growing tumors for which the first term in Eq. (1) dominates over the second. In this context, it will be assumed that *b* ≃ 0, meaning that most of the metabolic requirements are routed towards biosynthesis rather than basal energy consumption. Moreover, our dataset having time intervals with three consecutive longitudinal measurements without treament (*V*_0_, *V*_1_, *V*_2_) allows us to identify at most three parameters for each tumor (*V*_0_, *a, β*) but not more. When *b* is assumed to be zero, for a clonally homogeneous tumor with only a fraction of the cells proliferating due to necrosis, nutrient limitations, etc., exponents 0 < *β* < 1, correspond to growth slower than exponential, but with volumes still increasing with time.

Three patient groups were studied in a first batch of analyses. They included the cases of (i) growing untreated and (ii) relapsing post-radiotherapy BMs, and also (iii) growing BMs from patients under chemotherapy (CT) but with no specific treatment for the BMs. Patients in the last group included only drugs crossing the blood-brain-barrier (BBB) as described in *‘*Methods*’*. Figure 1(a,b) shows examples of BM longitudinal growth dynamics as observed in MRI studies. The growth exponent *β* governing the dynamic for each patient was obtained as described in the Methods section. The median value of the fitted individual exponents for untreated BMs (*N* = 10) was *β* = 1.59. This suggests substantial growth acceleration with *β* 3/2. It is interesting to note that this number differs from the value 5/4 obtained from metabolic scaling data of primary tumors [8].

**Figure 1.**
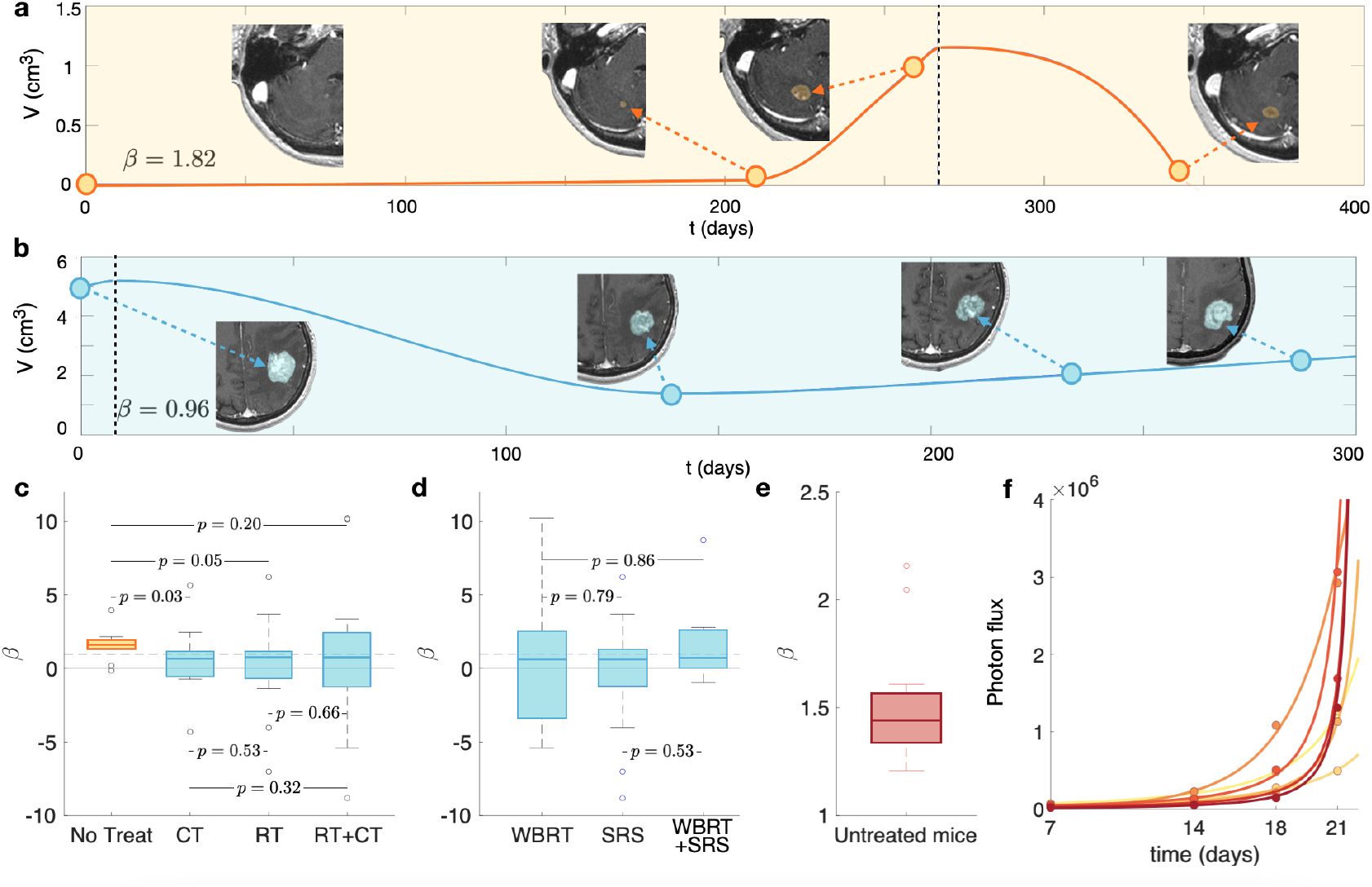
Growth dynamics of untreated and post-treatment relapsing BMs. Longitudinal dynamics observed in **a**. an untreated breast cancer BM and **b**. a relapsing post-SRS lung cancer BM. SRS treatment times are marked with a vertical dashed blue line. Yellow dots are the measured volumes and the solid blue lines are the result of interpolating longitudinal volumetric data with chirped cubic splines (shown only to guide the eye). Axial slices of the contrast-enhanced T1-weighted MRI sequences are displayed. **c**. Box plots comparing the growth exponents *β* of the different groups: untreated (*N* = 10), growing during chemotherapy treatment (CT, *N* = 16), recurrent BMs receiving only radiation therapy (RT, *N* = 23), or both (RT+CT, *N* = 33). Growth exponents were obtained for each BM using Eqs. 3. **d**. Box plots of the growth exponent *β* for BMs after RT: WBRT (*N* = 16), SRS (*N* = 31) and both (*N* = 9). The Kruskal-Wallis test gives non-signi*1*cant *p*-values, showing no differences between growth exponents *β* in these groups. **e**. Box plot of the growth exponents *β* in mice (*N* = 20) injected with an human lung adenocarcinoma brain tropic model (H2030-BrM). **f**. Total tumor mass growth curves for some of the studied mice where dots correspond to the measured values.

Median growth exponent for BMs growing under CT was *β* = 0.64 (*N* = 16). For BMs growing after completing radiation therapy (RT) we obtained *β* = 0.72 (*N* = 23). Finally, for those having received RT and under CT we got *β* = 0.68 (*N* = 33), where RT can stand for WBRT, SRS or a combination of both. Box plots are shown in Fig. 1(c). Similar growth exponents were obtained after receiving different RT modalities (see Fig. 1 (d)). Thus, volumetric growth of treated BMs had lower *β* exponents on average than those obtained for untreated BMs.

### Animal models recapitulating BM*’*s natural history display superexponential dynamics

To investigate the growth patterns of untreated BMs in faithful animal models, experiments in mouse models were performed as described in Methods. For the experiments, the human lung adenocarcinoma brain tropic model H2030-BrM3 was used since it is injected into the hearts of nude mice and leads to the formation of brain metastases from systemically disseminated cancer cells. Thus, the cell line used recapitulates at least in part the complex sequences of transformations required for cells to metastatize.

Since more than three points were available in the mice dataset, the growth exponents were computed using a different fitting technique as described in *‘*Methods*’*. The median value of the individual exponents *β* was 1.44 (*N* = 20) (Fig. 1 (e,f)).

### Sensitivity analysis of the exponents calculation

Since three points per patient were available to obtain three parameters, the fitting could be very sensitive to small variations in the data. Those variations in volumetric data could be given either by the time between MRIs or by image segmentation, regardless of being performed by the same image expert, and revised by another expert and a radiologist. In order to asses the effect of small changes in measured volumes on the results, an analysis of sensitivity was performed as explained in *‘*Methods*’*.

The growth exponent values were consistent in 74% of the BMs used in the study when a random error was added. Once sensitive cases were excluded, the results were in line with those of the entire cohort of BMs (Fig 2), ensuring the strength of the study.

**Figure 2.**
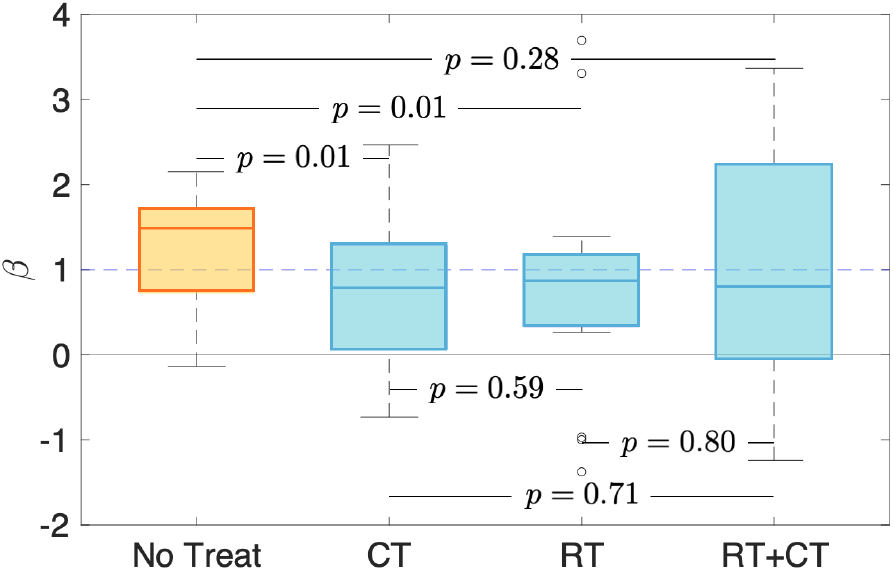
Growth dynamics of untreated and post-treatment relapsing BMs for the group with the most stable values of the growth exponent. Box plots comparing the growth exponents *β* of the different groups after a sensitivity analysis: untreated (*N* = 8), growing during chemotherapy treatment (CT, *N* = 14), recurrent BMs receiving only radiation therapy (RT, *N* = 18), or both (RT+CT, *N* = 21).

### Growth exponents best fitting the dynamics display super-exponential growth for untreated patients and subexponential growth for treated ones

An alternative approach to obtaining *β* was designed by looking for the value *β*_*_ that provided the best fit for all patients in each subgroup (untreated, growing while in CT treatment, and relapsing post-RT -with or without CT-RT*). The absolute errors weighted by volume, relative errors, were computed for each *β*_*_ value and each group of patients.

The exponent best fitting the whole dataset of untreated BMs was *β*_*_ = 1.5 (= 10). For treated relapsing tumors, the best fit was obtained with *β* = 0.51 (CT, = 16) and *β*_*_ = 0.71 (RT*, = 56), showing again a slowing down of the post-treatment growth dynamics (Fig. 3.b).

**Figure 3.**
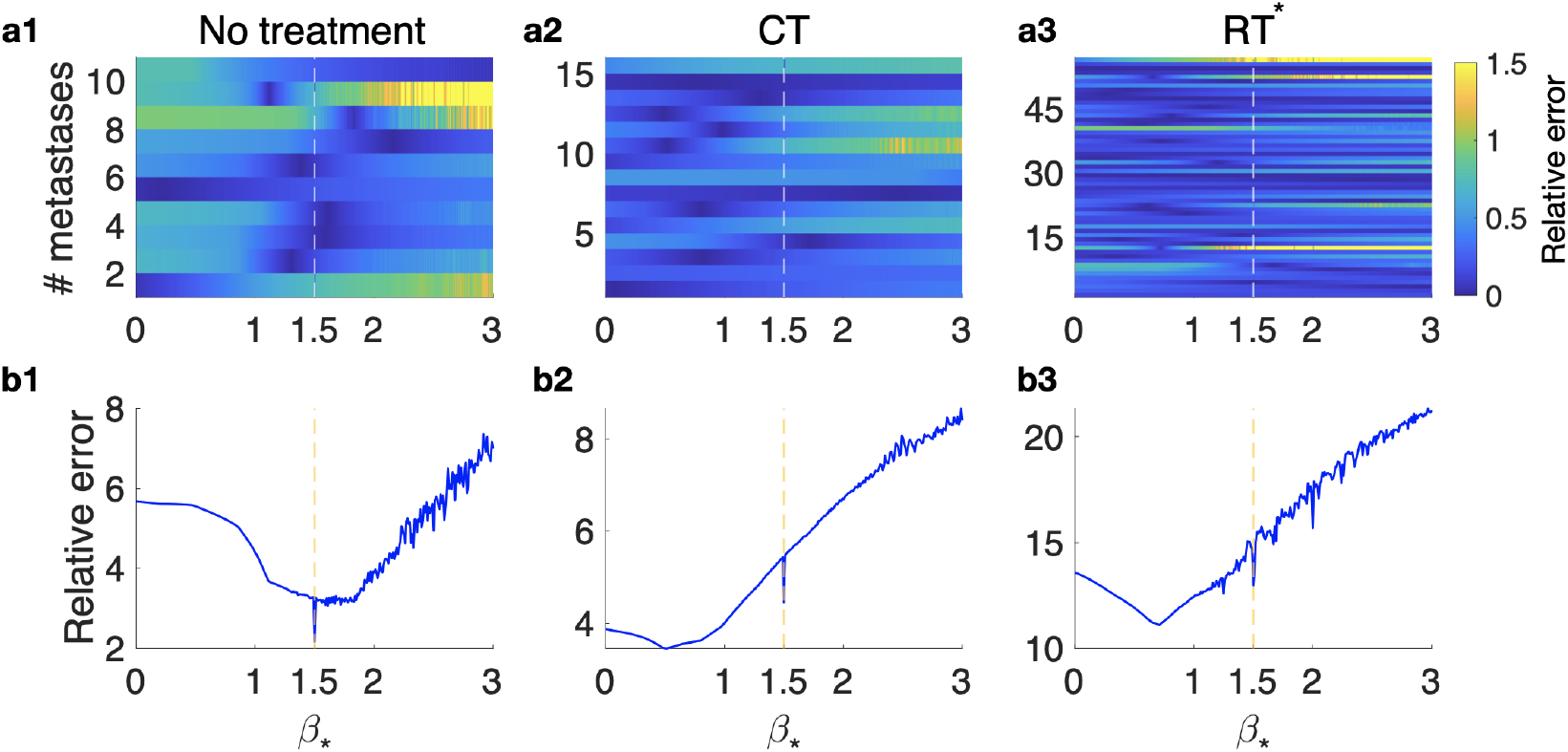
Growth exponent best *1*tting the dynamics for the dataset of untreated, CT and RT groups. Relative errors obtained when the *β* growth exponent is *1*xed. **a**. Errors computed for each metastasis in the different groups: untreated (**a1**), CT (**a2**), RT* (**a3**). **b**. Cumulative errors for each subgroup of BMs as a function of *β*_*_ for each of the different groups: untreated (**b1**), CT (**b2**), RT* (**b3**).

It*’*s interesting to note that at *β*_*_ = 1.5, a minimum (absolute or relative) appears in each dataset. Such minima are absolute for untreated lesions and relative in the case of treated ones, which indicates that after treatment, when the best growth exponent defining the group is lower than one, there is still a relevant tumoral component in some cases.

### Evolutionary dynamics of tumor complexity

It is known that cancer treatments lead to a reduction of the clonal complexity at the point of maximal response, due to the selective pressures exerted by the therapies [12–15]. Competition between clonal populations with different traits is a key ingredient leading to the increase of the growth exponent *β* [8]. Hence, one would expect this reduction in heterogeneity to be re*2*ected in the growth exponents of BMs as observed in our previous analysis.

The influence of different clonal compositions on the growth exponent of BMs was assessed *in silico* using an adapted version of the mesoscopic model developed in Ref. [16] (as described in Methods). We explored computationally a simplified scenario were BMs were made of two predominant clonal populations, one being more aggressive than the other. The initially most abundant population proliferated and migrated at fixed rates, while the least abundant population had an advantage in both processes, and hence was assumed to be more aggressive, due to either mutational changes or irreversible phenotype changes, providing evolutionary benefits. In that way we accounted for an initial tumor heterogeneity, and competition between the two populations was sustained during the early follow-up, several months post SRS.

First, the growth exponent *β* of an untreated virtual BM was evaluated. To do so, we simulated starting with a small fraction (10%) of an aggressive population coexisting with a larger population of less aggressive cells. After a few months, the tumor was substantially enriched *in silico* in the most aggressive population (94% versus 6%) and the growth exponent *β* was found to be *β* = 1.53 Fig. 4(a).

**Figure 4.**
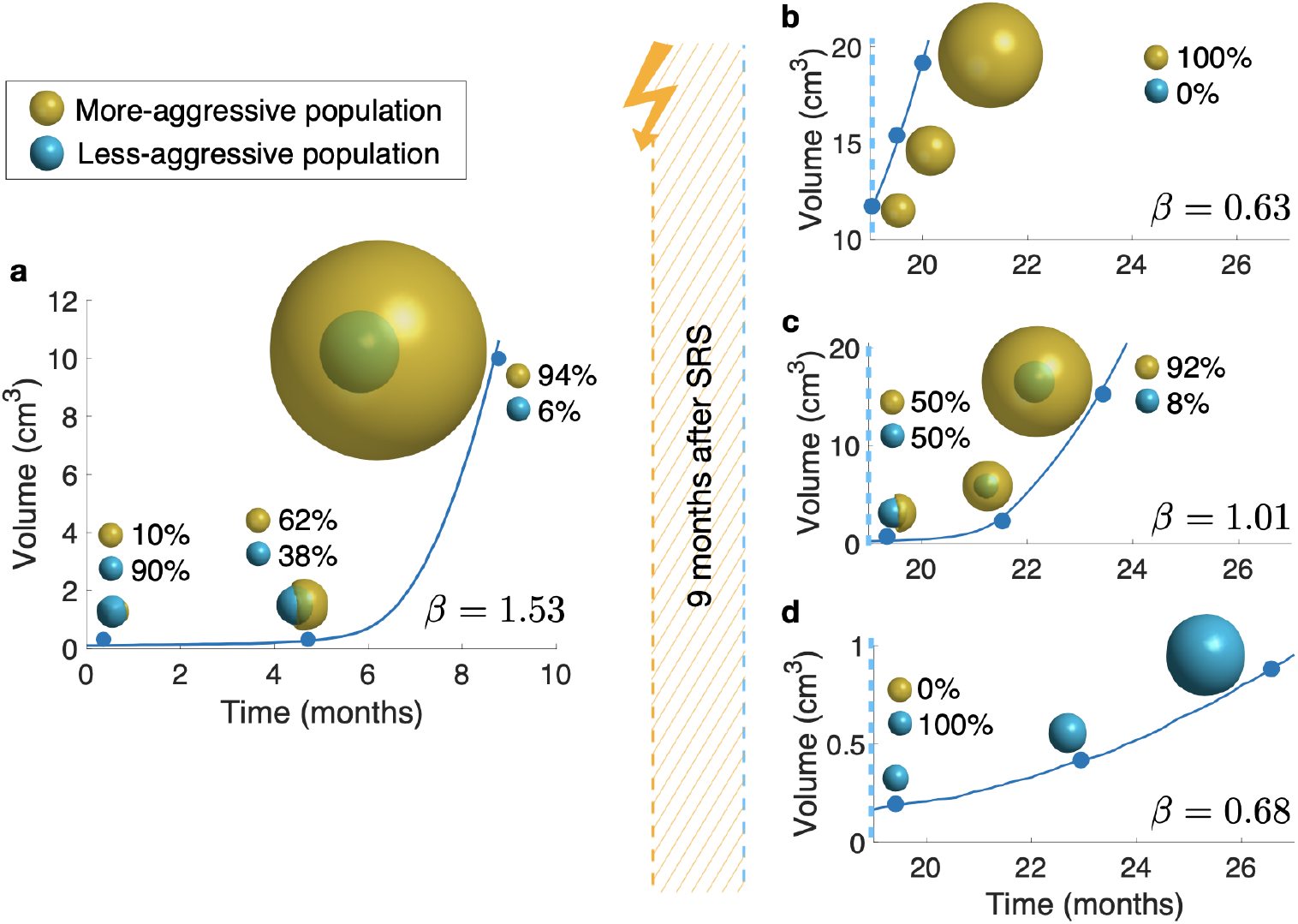
Simulations of longitudinal growth of heterogeneous BMs with two initial populations. (turquoise: less aggressive, and ocher: more aggressive). **a**. pre-treatment and **b**,**c**,**d** post-treatment cases.The more aggressive population carries an advantage of 80% in proliferation speed and 92.5% in migration speed, compared to the less aggressive population. In **a** the BM is composed of 10% of more aggressive cells, and 90% of less aggressive cells. After eight months, the more aggressive population has overcome its counterpart, becoming dominant. Then, three different situations that can happen after treatment are illustrated: **b**. the less aggressive population is completely removed from the tumor; **c**. both populations remain in a balanced state, and **d**. the more aggressive population is completely removed from the tumor. The betas were computed by choosing a random time point from each third of the total simulated time and are shown on each subplot.

In a second set of computational experiments, initially growing tumors were treated *in silico* with SRS with a differential effect on the two subpopulations. First, one or other of the subpopulations was assumed to be very sensitive to SRS, so that one of the populations was completely depleted. In both scenarios, depicted in Fig. 4(b,d) growth exponents were respectively *β* = 0.63 and *β* = 0.68, thus far from even exponential growth. A third, and more realistic, scenario assumed the most aggressive population to be more sensitive to treatment, thus restoring equilibrium between the two subpopulations. In this scenario, the growth exponent found was *β* = 1.01, still far from super-exponential, Fig. 4(c).

To study the influence of different advantages in proliferation and migration of the most aggressive population on the growth exponent *β*, simulations were performed varying the value of the advantage coefficients *v*_*div*_ and *v*_*mig*_ (see *‘*Methods*’*), and keeping the initial proportion of aggressive cells equal to 10%. We observed that the largest growth exponents *β* were obtained for combinations of slight advantages in both processes (*β* = 1.758 for *v*_*div*_ = 0.8, *v*_*mig*_ = 0.95; Fig. 5(b)) and that, for most combinations of coefficients *v*_*div*_ and *v*_*mig*_, the resulting exponent *β* is lower than 1.

**Figure 5.**
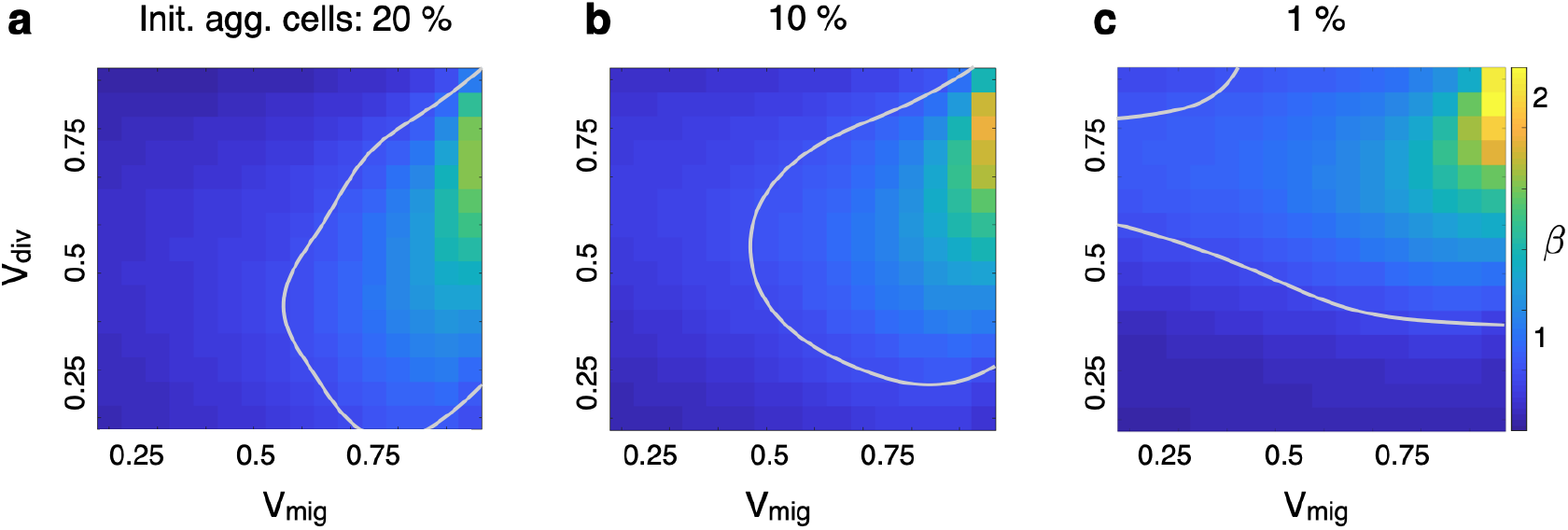
Growth exponents *β* obtained from a parameter sweep varying the advantages in migration and proliferation. Simulations were carried out for different combinations of coefficients *v*_*div*_ (advantage in proliferation; values explored range from 0.2 to 0.9) and *v*_*mig*_ (advantage in migration; values explored range from 0.2 to 0.95). **a** Poor-effectiveness treatment case (proportion of aggressive cells after treatment is 20%). The largest exponent *β* obtained is equal to 1.615, for a *v*_*div*_ = 0.75 and a *v*_*mig*_ = 0.95. **b** Medium-effectiveness treatment case (proportion of aggressive cells after treatment is 10%). The largest exponent *β* achieved was 1.758, for a *v*_*div*_ = 0.8 and a *v*_*mig*_ = 0.95. **c** High-effectiveness treatment case (proportion of aggressive cells after treatment is 1%). The largest exponent *β* achieved was equal to 1.99, for a *v*_*div*_ = 0.85 and a *v*_*mig*_ = 0.95. Gray lines correspond to *β* = 1.

Therapy is most harmful to the most proliferative cells [17, 18]. Hence, it is to be expected that the relative abundance of aggressive cells will decrease after treatment. To explore the influence of treatment on the growth exponent *β*, we repeated the previous analysis but this time the initial proportion of aggressive cells was modified to represent scenarios were the treatment was either very effective (initial proportion of aggressive cells is 1%, Fig. 5(c)) or not very effective (initial proportion of aggressive cells is 20%, Fig. 5(a)). The resulting *β* maps (each spot corresponds to a simulation performed with a fixed pair of coefficients *v*_*div*_ and *v*_*mig*_) are qualitatively similar, although it can be observed that the maximum exponent *β* achieved decreases as the initial proportion of aggressive cells increases. Again, we observed that most combinations of advantage coefficients yield a *β* lower than 1. This result suggests that, regardless of the resulting proportion of aggressive cells after treatment, it is rare to observe a growth exponent *β* larger than 1 in treated BMs.

## Discussion

Evolution is one of the main driving forces of life on Earth and is behind the observed diversity at every level of biological organization. Evolutionary processes are used by cancers to survive within their hosts and escape from the pressures exerted by treatments. It is a remarkable fact that the growth laws of untreated human malignant cancers and their animal model counterparts display a signature of the evolutionary processes taking place behind the scenes, in the form of an exponent *β >* 1 in (1).

This study mined a dataset including more than a thousand BMs to test such a surprising result over a time scale of months, i.e. the time interval spanning three MRI studies (6-9 months). BMs have a background of heterogeneity that could provide the necessary substrate for evolutionary competitive dynamics to happen, leading to super-exponential growth of the tumor mass. Phylogenetic analyses have revealed that BM-competent clones genetically diverge from their primary tumors at a relatively early stage in lung adenocarcinoma patients [19]. Genomic analyses of solid tumors and matched BMs revealed significant genetic heterogeneity between primary lesions and BMs [20], and the degree of genetic heterogeneity of BMs varied significantly among individuals with NSCLC, breast, and colorectal cancer [21–23]. In addition to the genetic heterogeneity, BMs have significant epigenetic variability [24, 25] and there can be other phenotype-based mechanisms playing a role [26].

Our results manifested both a macroscopic reflection of the evolutionary dynamics in the form of a *β >* 1 exponent for pre-treatment longitudinal dynamics, and also the loss of biological richness experienced by BMs after therapy, which lead to substantially reduced exponents *β*_*_ = 0.71 post-SRS. It has been hypothesized, using mathematical models, that treatment strategies in which an oncological *“*first strike*”* reduces the size and heterogeneity of the population, then followed by *“*second strikes*”* could lead to cancer extinction in metastatic disease [27, 28]. Our results show the effectiveness of the first-line SRS approach in providing an ecoevolutionary first-strike strategy for BMs. Independently of the volumetric reduction observed, which ends up being marginally irrelevant if the tumor recurs, the substantial reduction of the growth exponent implies a direct effect on the tumor ecological complexity. In the case of BMs, *“*second-strike*”* strategies could be provided by targeted therapies with better penetration than classical drugs across the blood-brain barrier, many of which are under investigation [29, 30]. It is also very interesting that information obtained from global macroscopic images could provide information on the underlying biological richness of these metastatic lesions. Thus *β* could be understood as an evolutionary exponent providing some information on the tumor heterogeneity.

It may seem naively counterintuitive that BMs subject to different treatment modalities (SRS, WBRT, CT) led to similar growth exponents *β*, since SRS is known to be substantially more effective than WBRT or CT. Our retrospective study focused only on BMs growing after (RT) or under (CT) treatment, but of course there would be many BMs with complete response, e.g. to SRS, that were not included here. It is also relevant to emphasize that the growth exponent cannot be directly interpreted as a growth rate, e.g. the speed of volumetric growth, but as a measure of the shape of the tumor growth curve.

An intriguing result of our study was the fact that the growth exponent for untreated BMs when fitted together was close to 3/2. It has recently been found for different primary tumors, lung (both adenocarcinoma and squamous cell), breast, colorectal, glioma, and head and neck cancer, that metabolic scaling exponents are close to 5/4 [8]. Following the classical reasoning of West et al. [31], one would expect metabolic exponents to be the same as growth exponents. This raises the interesting question of what the metabolic scaling of BMs would be on diagnosis, since the datasets of [8] did not include that condition. Would it be 3/2, raising the question of why BMs have a different metabolic scaling than other cancers? Or would it be 5/4, raising the question of why there should be a mismatch between metabolic and growth exponents in BMs? Data on metabolic scaling of BMs would be necessary to answer that question.

The *in silico* observation that the largest exponents *β* were achieved for combinations of slight advantages in both processes, division and migration, points out that a great advantage will not lead to a *β >* 1, as the overtaking time will be reached more quickly by the aggressive population, failing to sustain a competition throughout the time span of the BM. Another observation is that *β* decreases faster with changes along the *v*_*mig*_ axis, suggesting that advantages in migration may bring more competitive advantages in division.

Several authors have wondered whether the interval between SRS planning and treatment is accurate [32–35]. In order to study this, they compared volumes from diagnostic imaging and radiosurgery planning MRI and extrapolated the growth linearly to the day of SRS, since only two time points were available. Progression between diagnosis and SRS is common, and some suggest that a mathematical model would be useful to individualize treatments. Our model based on three time points shows that the growth velocity of untreated BMs increases with time. Thus pointing out an inadequate prediction of the tumor volume on treatment day and a substantial benefit of reducing the interval between SRS planning and treatment.

The main strength of our study was that it was based on a substantial dataset of 1133 BMs treated with SRS with high-resolution data, according to the guidelines for BM clinical studies [36]. Besides, each lesion was carefully segmented by the same expert and verified by a radiologist. Also it was a multicenter approach, including BMs from five different hospitals.

### Limitations of the study

The main limitation was that only a subset of 82 BMs of the whole 1133 BMs could be used for the computation of *β*. Only this part of the data had the three sequential imaging studies displaying sustained growth and with no further therapeutic actions performed in that time window required to compute the growth exponents. Because of the limited size of the final dataset, the data could not be analyzed separating patients by primary histologies. It would be very interesting to look at whether any of the conclusions of our study depend on the type of primary cancer considered.

### Conclusion

In summary, we studied a large BM dataset and unveiled a continuous acceleration of growth in the case of untreated lesions, due to Darwinian competition between different tumor subpopulations, as validated by in silico simulations using a stochastic discrete mesoscopic model. Results for mice data were in line with that. Recurrent BMs after treatment displayed slower growth, compatible with treatment-mediated reduction of tumor heterogeneity. As a result, we have highlighted the predictive value of a macroscopic variable, the evolutionary exponent, which can be used to obtain information on the microscopic status of the tumor.

## Data Availability

All data produced in the present study are available upon reasonable request to the authors.

## Author contributions

Conceptualization: BO-T, JP-B, EA, VMP-G. Methodology: BO-T, JJ-S, EA, JP-B, DM, VMP-G. Investigation: BO-T, JP-B, DM, JJ-S, DA, AO-M, BA, MV, LZ, PG-G, LP-R, VMP-G, EA, EG-P, MLl, NC. Software: BO-T, JP-B, JJ-S. Data curation: BO-T, JP-B, DM, VMP-G. Writing-Original draft: BO-T, VMP-G. Writing-Review and editing: BO-T, VMP-G, JP-B, JJ-S. Supervision: JP-B, DM, EA, VMP-G. Project administration: VMP-G. Funding acquisition: VMP-G.

## Supplementary

Supplementary material is available.

## Data availability

All data that support the plots within this paper and other findings of this study are available from the corresponding author upon reasonable request.

## Methods & Materials

### EXPERIMENTAL MODEL AND SUBJECT DETAILS

#### Patients

Patients included were all participants in the study MetMath (Metastasis and Mathematics), a retrospective, multicenter, nonrandomized study approved by five hospitals (blinded for review). All patients were diagnosed with BM in the period 2007-2021 and followed up with MRI according to standard clinical practice. A total of 354 patients who received SRS at any time during the evolution of the disease, and with full longitudinal follow-up, were reviewed in the study, including 1133 BMs. Primary tumor histologies were mainly non-small-cell lung cancer (NSCLC), breast cancer, melanoma and SCLC.

For the study of longitudinal volumetric dynamics, BMs were selected from the MetMath dataset on the basis of several inclusion criteria. First of all a minimum of three consecutive imaging studies, including a volumetric contrast-enhanced (CE) T1-weighted MRI sequence (slice thickness ≤ 2.00 mm, no gap) with no substantial imaging artifacts, at different time points, were required in order to allow for reliable lesion volume calculations. Secondly, an increase in tumor volume at each of the three time points was required, since it was desired to study the growth of either untreated or recurrent tumors avoiding. Next, only time points without previous SRS/WBRT treatments or with SRS/WBRT treatments received more than four months before the first imaging study were considered, in order to exclude the potential confounding effect of acute inflammatory responses seen in some patients in the first MRI after SRS. Patients with prior surgical resection of the metastasis were excluded to avoid confounding effects, such as ischemia. Brain metastases lacking relevant clinical variables and/or data on treatments received, as well as those lacking consensus in segmentations (what may lead to uncertain values of the volumes) were also excluded. Finally, patients were included regardless of their systemic treatment; however, BMs were considered as untreated for patients who were receiving chemotherapies (CTs) which are unable to cross the blood-brain-barrier (BBB) such as pertuzumab or trastuzumab [37, 38]. Those treatments target primary tumors but are known to have little or no effect on metastatic lesions due to the protecting effect of the BBB. Drugs known to cross the BBB were considered as treatments.

Each BM in our dataset was carefully revised to determine whether or not it satisfied the inclusion criteria. To do so lesion segmentation was necessary in many cases to assess its growth dynamics. Finally, 82 BMs from 58 patients were included in the study. Of these, 10 were untreated, 16 had received chemotherapy (CT), 23 WBRT or SRS (radiation therapy) and 33 received both treatments. A summary of patient characteristics used for the study is provided in Table 1.

**Table 1.** Summary of patient and BM characteristics, histology and volumetric parameters.

| Patient characteristics |  |
| --- | --- |
| Number of patients | 58 |
| Number of metastases | 82 |
| Age (years) | 58.48 (33-78) |
| Metastases per patient | 1.63 (1-19) |
| Sex (Male (M), Female (F)) | 53% M (31), 47% F (27) |
| Primary cancer Histology |  |
|  | Percentage (number of BMs) |
| NSCLC | 54.88% (45) |
| Breast | 26.83% (22) |
| Melanoma | 4.88% (4) |
| SCLC | 6.10% (5) |
| Others | 7.32% (6) |
| Volumetric parameters |  |
|  | mean (range) |
| Total tumor volume (cm<sup>3</sup>) | 3.45 (0.003-33.106) |
| CE volume (cm<sup>3</sup>) | 2.77 (0.003-28.989) |
| Necrotic volume (cm<sup>3</sup>) | 0.68 (0.000-12.452) |

#### Imaging and follow-up

The volumetric contrast-enhanced T1-weighted MR imaging sequence used to delineate the BMs and compute their volumes was gradient echo using 3D spoiled gradient-recalled echo or 3D fast-field echo after intravenous administration of a single dose of gadobenate dimeglumine (0.10 mmol/kg) with a 6-to 8-minute delay. MRI studies were performed in the axial or sagittal plane with a 1.0 T (= 5), 1.5 T (= 357) or 3.0 T (= 55) MR imaging unit. Imaging parameters were no gap, slice thickness of 0.52-2.0 mm (mean 1.3 mm), 0.4-1.1 mm (mean 0.5 mm) pixel-spacing and 0.4-2.0 mm sacing between slices (mean 1.0 mm).

Typical time spacing between MRI studies for BM follow-up was about 3 months for the institutions participating in the study. In our dataset, the median time between the first two MRI studies was 3.04 months while for the second it was 2.64 months.

#### Tumor segmentation

T1-weighted images were retrospectively analyzed by the same image expert (B.O.-T.) and reviewed by both an image expert with more than 6 years of expertise in tumor segmentation (J.P.-B., D.M.-G. or V.M.P.-G.) and a senior radiologist with 27 years of experience (E.A.). Segmentations were performed by importing the DICOM files into the scientific software package MATLAB (R2019b, The MathWorks, Inc., Natick, MA, USA). Each BM lesion was automatically delineated using a gray-level threshold chosen to identify the CE tumor volume. Segmentations were then corrected manually, slice by slice, using an in-house software as described in [39]. Necrotic tissue was defined as hypointense tumor regions inside CE tissue. CE and necrotic areas of the lesions were reconstructed, the tumor interfaces rendered in 3D. Tumor volume was computed as the volume within the surface delimiting CE areas.

#### Experiments in animal models

The human lung adenocarcinoma brain tropic model H2030-BrM3 (abbreviated as H2030-BrM) [40] was injected into the hearts of nude mice to induce the formation of brain metastasis from systemically disseminated cancer cells. H2030-BrM was cultured in an RPMI1640 medium supplemented with 10% FBS, 2 mM l-glutamine, 100 IU ml^−1^ penicillin-streptomycin and 1 mg ml^−1^ amphotericin B. Brain colonization and growth of metastasis were traced using non-invasive bioluminescence imaging, as BrM cells express luciferase. On administration of the substrate D-luciferin, bioluminescence generated by cancer cells was measured over the course of the disease. The increase in photon flux values is a well-established correlate of tumor growth in vivo [40]. The experiments were performed in accordance with a protocol approved by the Centro Nacional de Investigaciones Oncológicas (CNIO), the Instituto de Salud Carlos III and the Comunidad de Madrid Institutional Animal Care and Use Committee. Athymic nu/nu mice (Harlan) aged 4-6 weeks were used. Brain colonization assays were performed by the injection into the left ventricle of 100 *μ*l of PBS containing 100,000 cancer cells. Mice anaesthetized with isofluorane were injected retro-orbitally with D-luciferin (150 mg kg^−1^) and imaged with an IVIS Xenogen machine (Caliper Life Sciences). A bioluminescence analysis was performed using Living Image software (v.3).

#### Cell culture

H2030-BrM was cultured in an RPMI1640 medium supplemented with 10% FBS, 2 mM l-glutamine, 100 IU ml^−1^ penicillin-streptomycin and 1 mg ml^−1^ amphotericin B.

### MATHEMATICAL AND COMPUTATIONAL METHODS*’* DETAILS

#### VB growth model

Solving (1), with *b* = 0 leads to

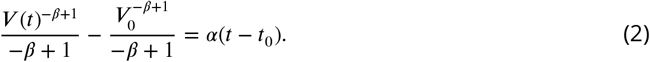

Since there is information about the dynamics at three time points (*t*_0_, *V*_0_), (*t*_1_, *V*_1_) and (*t*_2_, *V*_2_) obtained by image segmentation, the two parameters *α* and *β* can be completely determined by evaluating (2) at the times *t*_1_, *t*_2_, giving

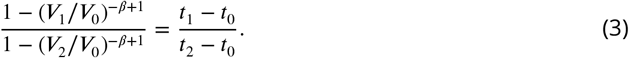

Eq. (3) is an algebraic equation for *β* that was solved using the MATLAB function fzero (which returns the root of a nonlinear function) for each set of known values *V*_0_, *V*_1_, *V*_2_, *t*_0_, *t*_1_, *t*_2_. A sensitivity analysis was performed to ensure the robustness of the method for the computation of *β*.

The growth exponent *β* provides information on the shape of the tumor growth curve, which cannot be directly interpreted as a growth rate, e.g. the speed of volumetric growth. Figure 6 (a) shows examples fitting the same pair of volumes and time points (initial and final), with different values of *β* while subfigure (b) shows that any fixed value of *β* (chosen there arbitrarily as *β* = 1, i.e. exponential growth), is compatible with different *‘*growth rates*’*.

**Figure 6.**
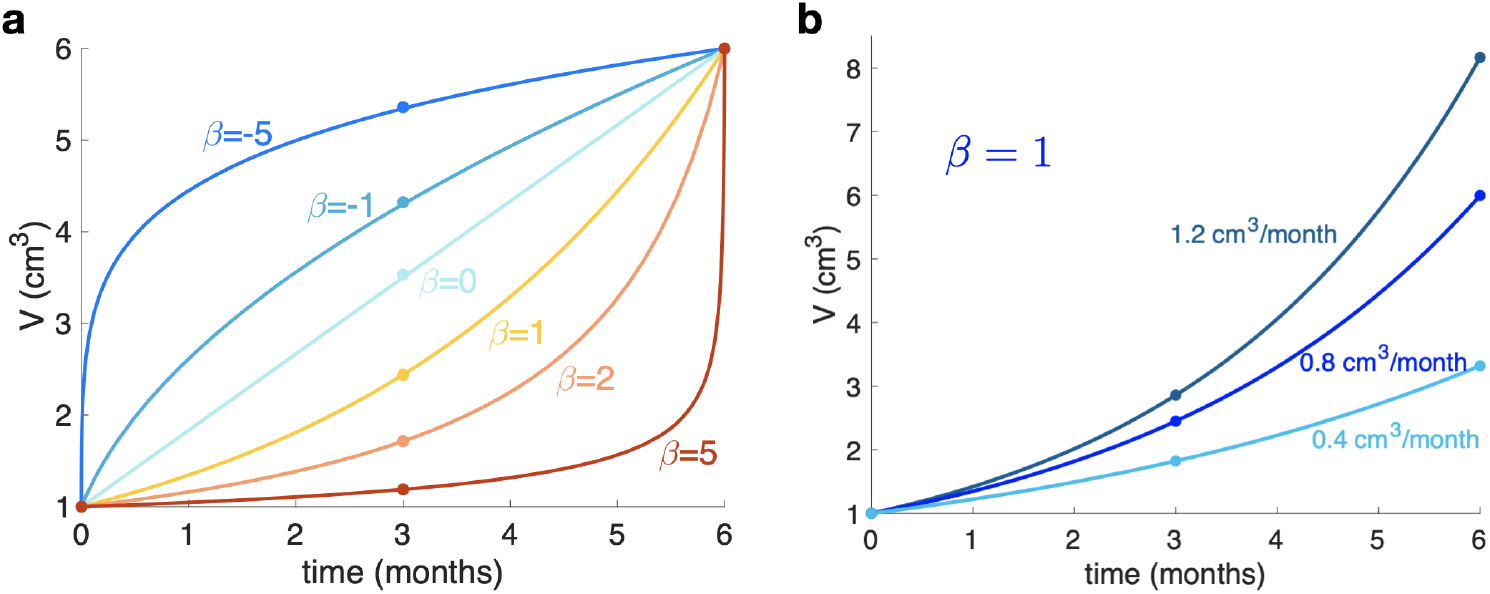
Interpretation of the growth exponent *β*. **a**. Growth behavior for several values of the growth exponent *β* when fixing initial and final volumes and times. The growth exponent gives information about the shape of the curves. **b**. Growth behavior when fixing *β* and the initial volume, showing different growth curves with different growth rates but the same exponent.

### Sensitivity analysis of the exponents calculation

To ensure the robustness of the growth exponent *β* computation, some well-known growth curves were studied: exponential, cubic, Gompertz and logistic. For each type of growth, 100 sets of points (*t*_1_, *t*_2_, *t*_3_), (*V*_1_, *V*_2_, *V*_3_) were randomly chosen and *β* was computed. For the exponential and cubic growths, the same value was obtained independently to the chosen points (Fig 7.b1-b2). In the case of Gompertz and logistic growths a set of *β* values was obtained (Fig 7.b3-b4). When adding a random error from -20% to +20% to the curves, a wider range of values for *β* appears, but such values are located around the values obtained without error (Fig 7.c).

**Figure 7.**
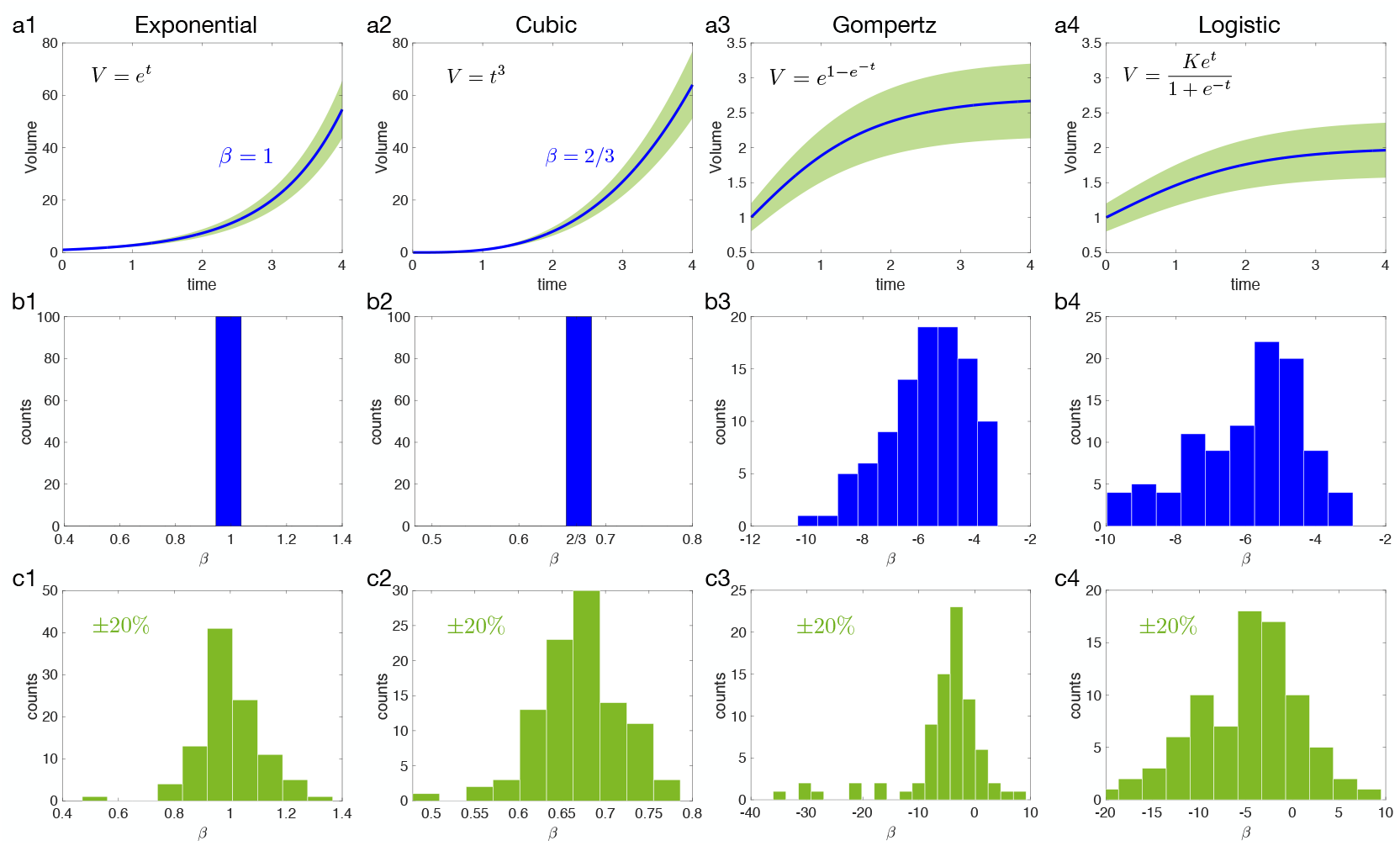
Growth exponent *β* for different types of growth: exponential, cubic, Gompertz and logistic. **a**. Growth function in blue with a ±20 % error in green. **b**. Computed *β* from Eq (1) for the different growth without error. **c**. Computed *β* when a random error is taken into account.

### Longitudinal growth analysis: individual growth exponents for mice

In animals, more than three volumetric points were available and a different method was used to calculate the growth exponent for each mouse. Measurements at 7, 14, 18, 21, 25 and 28 days were usable from 24 mice, however at 25 and 28 days, the measured volume was close to the total brain volume and it was assumed that growth would be affected by limitations of space. Thus, to avoid confounding effects coming from mechanical constraints, the latter two time points were excluded from the analysis. A discretization of Eq. (1) was performed, and taking logarithms on both sides

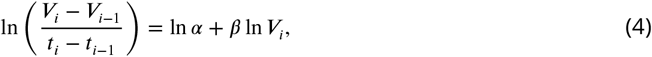

the slope of the straight line that best fits all the points for each mouse corresponds to the growth exponent *β*. Four mice were excluded because of volume decrease, leading to the use of 20 mice for *β* computation.

### Sensitivity to small changes in volume when computing beta

In order to test the sensitivity to little fluctuations in the data, a random error smaller or equal to ±5% was added to each volume. For each BM, the procedure was carried out 200 times in order to compute *β*\* for each set of random errors. It was imposed that the average of the 200 calculated *β*\* have a difference less than 0.5 from the computed *β* for the measure volumes, that is to say,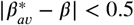.

### Longitudinal growth exponents: Group calculations

An iterative method was used to automatically compute the optimum *β*_*_, that is, the one giving the lowest relative error to the segmented volumes for each group. A sweep was performed on *β* = [0, 3] with 300 steps and on 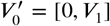 and 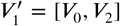, with 500 steps for each. To each *β* and each pair 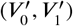 there corresponds a single value of 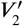 from Eq. (3). Then, the sum of relative errors of the three segmented volumes for every BM (*V*_0_, *V*_1_, *V*_2_) was computed using the formula:

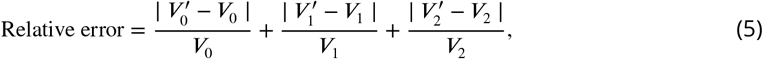

the smallest is retained, that is to say, the combination of 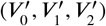 that best fits (*V*_0_, *V*_1_, *V*_2_), given each *β* (pseudocolor plots in Fig. 3(a).) Finally, the *β*_*_ value for which the sum of all errors (Fig. 3(b)) is minimum and therefore corresponds to the best fit for the whole subgroup, is observed.

### Stochastic discrete mesoscopic simulator of BMs longitudinal dynamics and response to treatments

To illustrate how treatment could modulate heterogeneity and influence the *β* values obtained from BM longitudinal growth data, as well as to to provide an *in silico* testbed capable of simulating the growth and evolution of BMs, an adapted version of the mesoscopic model developed in [16] was used. The mesoscopic model is a discrete stochastic simulator of tumor growth, that features clonal populations as the basic agent, instead of individual cells. The spatial domain is divided into voxels, a 3D generalization of a bi-dimensional compartment (commonly used in medical imaging). Each voxel has a given carrying capacity *K*, so it can harbor as much as *K* cells, that may belong to different clonal populations. It is assumed that cells belonging to the same clonal population and exposed to the same stimuli (microenvironment, surrounding cell density, nutrient/oxygen availability) will behave in the same way. Therefore, instead of assessing cell-by-cell the outcome of any dichotomous success/failure process that an individual cell may perform (such as division or death), the number of cells in each clonal population that succeed in performing such processes is assessed in a single step.

If the outcome of each individual process *i* performed by a cell is considered as a random variable following a Bernoulli distribution *X*_*i*_ ∼ Bernoulli(*P*_*i*_), with a probability *P*_*i*_ associated to that process, then the joint outcome of an entire clonal population of *N* identical cells performing such process can be considered as a random variable following a binomial distribution 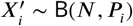. In this way, by knowing the number of cells *N* attempting to perform a process *i*, and the probability *P*_*i*_ of successfully performing that process, updating the number of cells in a clonal population is done by sampling the binomial distribution associated with that process.

The processes considered in this version of the model are division, death and migration. Note that mutations or phenotypic transitions are disregarded, as cells are not allowed to change from a given population to another. This assumption is grounded in the short time scale of evolution of BMs. At each time step, the cell number in each voxel is updated in a synchronous way (using swapping matrices) by random sampling the binomial distributions corresponding to each process, voxel and clonal population. The probabilities associated with these binomial distributions were defined in the same way as in the original work [16]. The version of the model used in this paper is adapted to simulate BMs. It considers only two clonal populations, with different traits and characteristic rates. The rates of division, death and migration were fitted (using ABC rejection algorithm [41]) to mimic the lifespan and volume dynamics of an actual BM.

One of these clonal populations is more aggressive than the other; this is implemented by faster division and migration rates. Namely, the advantage associated with the division process will be *v*_*div*_ E (0, 1], while the advantage associated with the migration process will be *v*_*mig*_ E (0, 1]. These coefficients represent the ratio between the characteristic time in which the aggressive cells carry out the considered process, versus the characteristic time of the other cell type. Hence, the lower their value, the greater the advantage associated with the aggressive population. The range of parameter values used in the model to perform simulations can be seen in Table 2. A cohort of 720 simulations was produced for the parameter exploration performed in Fig. 5.

**Table 2.** Summary of the most relevant parameters used for simulations of the BM mesoscopic model.

| Spatial and time domain parameters |  |  |
| --- | --- | --- |
| Time step length ($\Delta t$) | 4 | hours |
| Number of voxels per dimension | 80 |  |
| Voxel size | 1 | mm |
| Voxel volume | 1 | mm<sup>3</sup> |
| Carrying capacity ($K$) | $2 \cdot 10^5$ | mm<sup>3</sup> |
| Initial and final conditions for simulation |  |  |
| Initial cell number | 10 |  |
| Limit volume of virtual BM | 10 | cm<sup>3</sup> |
| Characteristic rates of each cellular process |  |  |
| Division rate | 0.36 | days<sup>-1</sup> |
| Death rate | 0.144 | days<sup>-1</sup> |
| Migration speed | 2.88 | mm<sup>2</sup> days<sup>-1</sup> |
| Advantage coefficients |  |  |
| Advantage in division ($v_{div}$) | 0.2-0.9 | a.d. |
| Advantage in migration ($v_{mig}$) | 0.2-0.95 | a.d. |

The adapted version of the mesoscopic model for BMs was coded in Julia (v. 1.1.1). Data processing and visualization of simulation files was performed in Matlab (v. 2018b).

## QUANTIFICATION AND STATISTICAL ANALYSIS

### Statistical analyses

Statistical analyses were performed using the MATLAB software, and also SPSS (Statistical package for the Social Sciences, v24.00 IBM) software. The normality of the variables was assessed via the Kolmogorov-Smirnov test. The Kruskal-Wallis test was conducted with adjustment for multiple comparisons, to determine statistically significant differences for non-parametric data (the scaling law growth factor, *β*). P-values smaller than 0.05 were considered to be statistically significant.

